# Genetic Variations in Evolutionary Accelerated Regions Disrupt Cognition in Schizophrenia

**DOI:** 10.1101/2021.02.24.21252224

**Authors:** Upasana Bhattacharyya, Prachi Kukshal, Triptish Bhatia, Smita N Deshpande, B.K Thelma

## Abstract

Cognition is believed to be a product of human evolution, while schizophrenia is ascribed as the by-product with cognitive impairment as it’s genetically mediated endophenotype. Genomic loci associated with these traits are enriched with recent evolutionary markers such as Human accelerated regions (HARs). HARs are markedly different in humans since their divergence with chimpanzees and mostly regulate gene expression by binding to transcription factors and/or modulating chromatin interactions. We hypothesize that variants within HARs may alter such functions and thus contribute to disease pathogenesis. 49 systematically prioritized variants from 2737 genome-wide HARs were genotyped in a north-Indian schizophrenia cohort (331 cases, 235 controls). Six variants were significantly associated with cognitive impairment in schizophrenia, thirteen with general cognition in healthy individuals. These variants were mapped to 122 genes; predicted to alter 70 transcription factors binding sites and overlapped with promoters, enhancers and/or repressors. These genes and TFs are implicated in neurocognitive phenotypes, autism, schizophrenia and bipolar disorders; a few are targets of common or repurposable antipsychotics suggesting their draggability; and enriched for immune response and brain developmental pathways. Immune response has been more strongly targeted by natural selection during human evolution and has a prominent role in neurodevelopment. Thus, its disruption may have deleterious consequences for neuronal and cognitive functions. Importantly, among the 15 associated SNPs, 12 showed association in several independent GWASs of different neurocognitive functions. Further analysis of HARs may be valuable to understand their role in cognition biology and identify improved therapeutics for schizophrenia.

## Introduction

The complexity of the human brain plays a crucial role in developing improved cognitive functioning and has led to distinct patterns of behavior in humans compared to their primate relatives (Heyes 2012; Srinivasan et al. 2018). During the course of evolution, humans have developed complex language (d’Errico et al. 2003), executive functioning and abstract thinking (Barkley 2001). This derives support from different patterns of performance witnessed across cognitive domains by humans and chimpanzees (Tomasello and Herrmann 2010). Thus, higher cognitive functions are regarded as one of the main distinctive traits of humans (Maclean 2016) and is considered to be a product of human evolution. On the contrary, schizophrenia has been hypothesized to be a by-product of recent evolution of the human genome from its closest ancestor, the chimpanzee (Liu et al. 2019; Avila, Thaker, and Adami 2001; T. J. Crow 2000; Timothy J. Crow 1997; Srinivasan et al. 2016; Van Dongen and Boomsma, n.d.).

Deficits in cognitive ability is a signature of schizophrenia. This trait has remained as the most valuable predictor of functional outcome over other symptoms (Rund 1998; Green et al. 2000) with more than 80% of patients showing significant cognitive impairment(Emre, Murat, and Christos 2010). Along with deficits in global cognitive ability, specific domains such as working memory, attention and executive function are also impaired in schizophrenia patients and like the primary disease pathology, these features are also highly heritable (Calafato and Bramon 2019) and clearly meet the criteria for being classified as genetically mediated endophenotypes (Harvey, n.d.). Genome Wide Association Studies (GWASs) of cognitive traits such as intelligence, general cognition (g-factor), reaction time, verbal-numerical reasoning, educational attainment (a proxy measure of cognitive function) have revealed overlaps in genetic risk loci between schizophrenia and cognitive traits (Hubbard et al. 2016; Smeland and Andreassen 2018; Le Hellard et al. 2016). Neurocognitive impairment being a core symptom of schizophrenia and resistant to first-generation antipsychotics and shows only a little improvement with second-generation antipsychotics (Ohi et al. 2018; Bowie and Harvey 2006), uncovering its genetic underpinnings and consequently biology with possibly new therapeutic options is an unmet need. It has been suggested that the genomic regions which are advantageous for the acquisition of important human traits such as language, complex cognitive skills and other favorable brain functions including behavioral flexibility also predispose to schizophrenia are also (Srinivasan et al. 2016). Enrichment of recent evolutionary markers in the vicinity of schizophrenia (Xu et al. 2015; Srinivasan et al. 2016) and cognition (Srinivasan et al. 2018) associated loci, suggest their involvement and importance for both these traits. Human accelerated regions (HARs) are one such class of genetic markers of recent evolution in humans.

HARs are segments of DNA that are well conserved throughout vertebrate evolution but are strikingly different in humans since their divergence from chimpanzees (Pollard et al. 2006). Genes associated with these HARs have been linked to expansion and function of higher-order cognitive networks in the human brain (Wei et al. 2019), human cognitive functioning, social behavior, brain development (Bae, Jayaraman, and Walsh 2015) and to brain disorders related to cognitive impairment such as autism spectrum disorder (Doan et al. 2016) and schizophrenia(Xu et al. 2015). Furthermore, among all the approved drugs, 32.42% target at least one HAR gene and notably, a significant proportion of these HAR-gene targeting drugs are used for treatment of neurological disorders implying their relevance in drug repurposing as well as in new therapeutics (Chu, Quan, and Zhang 2020). HARs are mostly intergenic (60.7%) or intronic (32.4%) in their distribution (Doan et al. 2016). Changes in protein coding sequences have primarily contributed to evolution across related species, however, large phenotypic divergence between humans and chimpanzees was probably primarily driven by changes in gene regulation (King and Wilson 1975; Carroll 2005). In accordance with this, HARs have been found to regulate expression of nearby genes by acting as enhancers, repressors, modifiers etc (Capra, Erwin, Mckinsey, et al. 2013; Ryu et al. n.d.) and induce human-specific gene expression (Kamm et al. 2013; Hubisz and Pollard 2014; Capra, Erwin, McKinsey, et al. 2013a; Prabhakar et al. 2008). Furthermore, a comparison between human and chimp brains showed human specific brain connections have significantly higher expression of HAR genes in cortical areas of the human brain (Van Den Heuvel et al. 2019). These findings suggest that HARs represent a modest portion of the evolutionary aspects witnessed in humans and could be functionally relevant for human specific development.

Based on these findings, we hypothesize that expression of genes in the vicinity of HARs could be altered by genetic variants present within the HARs either through alteration of the binding sites of transcription factors and/or other regulatory proteins; and/or their own epigenetic signature(s). Thus, identification of HAR variants associated with cognitive impairment in schizophrenia patients may enhance our understanding of the genetics and biology of cognitive impairment and may also help in identifying new drug targets.

## Materials and Methods

### Recruitment and diagnostic assessment of patients with schizophrenia

Inclusion criteria for recruitment of patients with schizophrenia and healthy controls, predominantly of north Indian origin have been previously described(Kukshal et al. 2013a). Briefly, with approval from the Institutional Ethics Committees of all of the participating institutions, patients diagnosed with schizophrenia as per Diagnostic and Statistical Manual of Mental Disorders, Fourth Edition (DSM-IV), were recruited either with their written informed consent or from accompanying relatives or caregivers, at the Post Graduate Institute of Medical Education and Research (PGIMER) -Dr. Ram Manohar Lohia (RML) Hospital, New Delhi. Hindi version of the Diagnostic Interview for Genetic Studies (DIGS) and the Family Interview for Genetic Studies (FIGS)(Deshpande et al. 1998; Nurnberger et al. 1994) were used to assess all the participants. Healthy adult controls in the study were recruited from matched population with their written consent, also described previously(Kukshal et al. 2013a).

### Neurocognitive assessment

Cognitive assessment of the participants recruited as above was carried out using the Hindi version of University of Pennsylvania Computerized Neurocognitive Battery (Penn CNB) as described in our previous studies (Bhatia et al. 2012; Kukshal et al. 2013b). The Penn CNB assesses eight cognitive domains commonly reported to be impaired in patients with schizophrenia, namely: abstract and mental flexibility, attention, face memory, spatial memory, working memory, sensory-motor processing, and emotional processing. The normalized Z scores were extracted from the data repository at CNB site at University of Pennsylvania for the eight selected cognitive domains as mentioned previously (Bhatia et al. 2012; Kukshal et al. 2013b). Briefly, three performance functions, namely accuracy, speed and efficiency were calculated for each of the eight domains. These scores were then used for association testing.

### Selection of variants from HARs

A consolidated list of all HARs (2737) reported in a recent study on role of HARs in autism(Doan et al. 2016) has been used in this study. Selection criteria for HAR-SNPs to be genotyped, have been described by us previously (Bhattacharyya et al. 2020). Briefly, from a total of 30,045 SNPs from within these HARs (hg19; dbSNP146 module in UCSC table browser; https://genome.ucsc.edu/) 2034 variants were prioritized based on minor allele frequency (MAF) [0.4< (MAF) > 0.1] in south Asian population. Based on RegulomeDB(A. P. Boyle et al. 2012) score (<3) and/or significant eQTLs in different regions of the brain, a total of 49 SNPs (Supplementary Methods, Supplementary Table 1) were further prioritized for genotyping in the north Indian case-control cohort recruited as detailed above,

### Genotyping

SNP Type Assays are allele-specific PCR assays designed by Fluidigm, which use three primer pairs and two universal probes to differentiate between the two alleles. Nanofluidics based Fluidigm SNP Type(TM) genotyping assay was performed at a commercial facility (Sandor Lifesciences Pvt. Ltd, Hyderabad, India) to genotype the 49 SNPs.

### Statistical Analysis

#### Data Transformation

Cognition scores, if not normally distributed, were transformed by skew power transformation [z = zbcn,u(y, λ, γ)](Hawkins and Weisberg n.d.) in cases and controls separately using car package in R(Fox and Weisberg n.d.) as described previously (Punchaichira et al. 2020).

#### Principal Component Analysis

The Kaiser-Meyer-Olkin (KMO) sample adequacy test and Bartlett’s test of sphericity were performed to determine the effectiveness of the PCA for neurocognitive measures. A Scree Plot of Eigenvalue versus component number was produced to determine the number of components explaining the greatest amount of variance in the traits. PCA was performed using the Direct Oblimin rotation method to reduce neurocognitive measures across eight domains to their principal components.

#### Linear Regression

A two-way ANCOVA was performed with health status and genotypes as fixed factors and each principal component of accuracy, processing speed and efficiency as dependable variables with age and gender as covariates using IBM® SPSS® Statistics (Version 27) (IBM Corp. Armonk, New York USA) as previously done in a similar study(Hori et al. 2012). One-way ANCOVA with age and gender as covariates separately for control groups were performed to further explore the simple effect of genotype on cognition.

### Functional annotation

Significantly associated SNPs following the regression analysis were functionally annotated using FeatSNP(Ma et al. 2019) (http://featsnp.org) as previously described(Bhattacharyya et al. 2020) (Supplementary Methods).

### Mapping HAR SNPs to genes

FUMA-SNP2GENE module was used to map SNPs to genes using three approaches have been used for mapping: (i) Positional, (ii) eQTL and (iii) Chromatin interaction as previously described(Bhattacharyya et al. 2020) (Supplementary Methods).

### Pathway enrichment and functional annotation of the mapped genes and transcription factors

Enrichment analysis was done using EnrichR (Chen et al. 2013; Kuleshov MV 2016) which is an intuitive enrichment analysis web-based tool providing various types of visualization summaries of collective functions of gene lists using 180,184 annotated gene sets from 102 gene set libraries such as KEGG, Reactome, NCI-Nature etc. VarElect was used to identify if the mapped genes were previously implicated in cognition. This is a tool for disease/phenotype-dependent gene/variant prioritization which uses different databases such as GeneCards®, MalaCards, LifeMap Discovery®, and PathCards. VarElect acts jointly on the gene list and phenotype/disease keywords, and produces a list of prioritized, scored, and contextually annotated genes and direct links to supporting evidence and further information. Details of the scoring method and search terms used by VarElect is provided in supplementary method. Furthermore, attempts were made to confirm if, the gene-sets identified in this study are associated with similar Human Phenotype Ontology (HPO)-Phenotypes. Statistical significance of these overlaps, if any, were tested using hypergeometric test (http://nemates.org/MA/progs/overlap_stats_prog.html). Basic equation to obtain the probability score of an overlap of genes using the above-mentioned program is provided in supplementary methods.

### Status of the fifteen associated SNPs in other studies

We checked for the association status of the all the 15 SNPs, in the publicly available GWAS data with large sample size on cognition, education attainment, neuroticism, intelligence. Details of these GWASs are provided in Supplementary Table 2.

## RESULTS

### Demographic details

A total of 331 schizophrenia cases and 235 healthy controls from north Indian population, were successfully enrolled in this study. Demographic data of the study cohort is shown in Table 1.

**Table 1:** Demographic details of the study cohort.

| | Cases (mean age±SD) | Controls (mean age±SD) |
| --- | --- | --- |
| Male | 231 (34±10) | 142 (40±16) |
| Female | 99 (32±9) | 93 (35±11) |
| Total | 331 | 235 |

### Data transformation and Principal component analysis (PCA)

Cognition scores that were not normally distributed, were transformed by skew power transformation and then PCA was conducted on the converted neurocognitive measures of all the eight domains with oblique rotation (Direct Oblimin). Verification of the sampling adequacy for the analysis was done with Kaiser–Meyer–Olkin (KMO) measure. The KMO value for cognitive scores of the eight domains was ≥0.770, i.e, above the acceptable score of 0.5. Bartlett’s test of sphericity was [χ2(276)=16788.815, p<0.001] indicated a high correlation between cognitive scores and thus requiring a PCA. Eigenvalues were obtained for each component by running an initial analysis on data. First six components had eigenvalues>1. By observing the scree plot (Figure 1) and the variance explained by the components (Supplementary Table 3), first two components namely component 1 and 2 were selected for two-way ANCOVA. The factor loadings after rotation are given in Supplementary Table 4.

**Figure 1:**
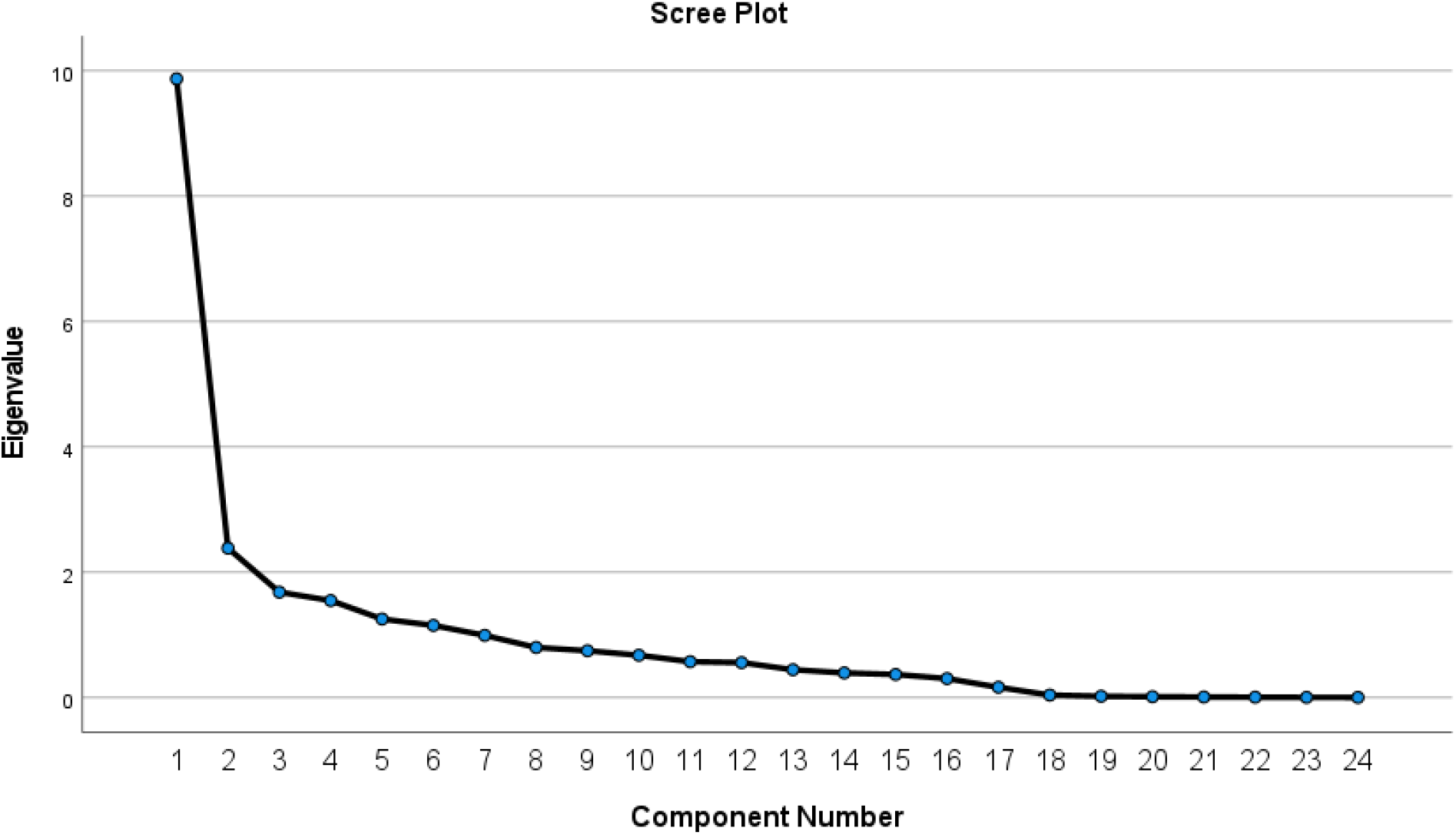
Scree plot.

### Status of the fifteen associated HAR SNPs in related GWASs

The association status of the 15 associated SNPs (13 in control group, 6 in case-control cohort, four common) in related GWASs such as general cognitive function(Gail Davies et al. 2018; G. Davies et al. 2016; Lee et al. 2018), educational attainment(Lee et al. 2018; Okbay, Beauchamp, et al. 2016), neuroticism(Mats Nagel, Jansen, et al. 2018; Okbay, Baselmans, et al. 2016; M. Nagel et al. 2020; Luciano et al. 2018; Turley et al. 2018; Mats Nagel, Watanabe, et al. 2018) and intelligence(Sniekers et al. 2017; Savage et al. 2018) were documented. As all the SNPs are intronic/intergenic and they were functionally annotated to understand their regulatory roles, if any. As mentioned before, HARs regulate genes around them and SNPs within these regions which may affect such regulatory functions might interfere with the expression of genes under their control. To further elucidate the role of HARs in cognition, identification of genes that may be impacted by one or more these 15 variants, as well as the pathways to which they may contribute was undertaken.

Of the 15 SNPs screened for in such GWAS datasets, all but two SNPs (rs214304, rs492804) showed nominal association (P-value<0.05) for with one or more related phenotypes such as general cognitive function, reaction time, verbal and number reasoning, memory and education attainment and intelligence. Most importantly, among these SNPs, rs1868172 was significantly associated with intelligence (P-value<2.33E-05). (Supplementary Table 5). Of note, though our sample size is low to modest, we obtained significant leads from this study.

### Functional annotation of associated SNPs

Function of the 15 associated SNPs was then annotated using FeatSNP (Table 3, Supplementary Table 6). These 15 SNPs were found to modulate the binding sites of 79 transcription factors based on transcription factor binding motif scores as calculated by FeatSNP. In addition, a few SNPs (rs214304, rs995089, rs1868172, rs11631291, rs6082296) were also found to overlap with histone ChIP-seq peaks that have methylation marks for active promoters, repressors or enhancers such as H3K4me3, H3K4me1, H3K9ac, H3K27ac, H3K36me3, H3K27me3, H3K9me3 (Table 3, Supplementary Table 6).

**Table 2:** Effect of genotypes and disease status on neurocognitive scores and effect of genotypes on neurocognitive scores of control group with age and gender as covariates.

| rs ID (Chromosome:bp;Effect allele) | A) Main effect of SNPs on cognition in control group (one way ANCOVA) |  |  |  | B) Effect of SNPs and disease phenotype on cognition (two way ANCOVA) |  |  |  |
| --- | --- | --- | --- | --- | --- | --- | --- | --- |
|  | Principle component 1 (2,15) |  | Principle component 2 (2,15) |  | Principle component 1 (4,170) |  | Principle component 2 (4,170) |  |
|  | F | P-value | F | P value | F | P-value | F | P value |
| rs214304 (1:19688164;C) |  |  | 4.03 | 0.04 |  |  |  |  |
| rs995089 (1:45098461;T) | 3.75 | 0.04 |  |  |  |  |  |  |
| rs3113980 (1:75160365;T) | 5.63 | 0.01 |  |  | 3.3 | 0.01 |  |  |
| rs492804 (1:170612262;T) | 4.33 | 0.03 |  |  |  |  |  |  |
| rs1868172 (3:139493701;T) | 6.68 | 0.008 |  |  |  |  |  |  |
| rs13121833 (4:1357358;C) | 4.71 | 0.02 |  |  | 2.54 | 0.04 |  |  |
| rs2659510 (4:102113048;C) | 7.28 | 0.006 |  |  |  |  |  |  |
| rs26719 (5:92136381;T) |  |  | 4.26 | 0.03 |  |  |  |  |
| rs36039219 (7:11704538;A) | 4.44 | 0.03 |  |  |  |  |  |  |
| rs10810957 (9:18332000;C) |  |  |  |  |  |  | 2.23 | 0.035 |
| rs11631291 (15:71889320;A) |  |  |  |  | 2.47 | 0.04 |  |  |
| rs12905641 (15:78964362;T) | 5.34 | 0.01 |  |  |  |  |  |  |
| rs1885986 (17:2203175;G) | 4.81 | 0.02 |  |  | 2.57 | 0.04 |  |  |
| rs10423094 (19:49533380;T) |  |  | 3.92 | 0.04 |  |  |  |  |
| rs6082296 (20:21026509;C) | 4.71 | 0.02 |  |  | 3.08 | 0.01 |  |  |

**Table 3:** Functional annotation of associated SNPs.

| rs ID<br>(Chromosome:bp;Effect<br>allele) | Transcription Factor motif predicted to be interfered | Overlap with histone<br>ChIP-seq peaks in brain<br>tissue# |
| --- | --- | --- |
| rs214304 (1:19688164;C) | *MAFG:NEF2L1, MEIS1, MEIS3, MEOX1, MEOX2* | H3K4me1 |
| rs995089 (1:45098461;T) | *NKX2-5* | H3K4me3, H3K4me1,<br>H3K9ac, H3K27ac,<br>H3K36me3 |
| rs3113980 (1:75160365;T) | *FOXD1* | — |
| rs492804 (1:170612262;T) | *FOXD1, FOXL1, FOXQ1, FOXJ2, FOXO4, FOXO6, FOXP3,*<br>*FOXK1, FOXG1, FOXQ1, FOXI1* | — |
| rs1868172<br>(3:139493701;T) | *FOXD1, FOXF1, FOXG1, FOXJ3, FOXK1, FOXO3, FOXO4,*<br>*FOXO6, FOXP3* | H3K4me1, H3K27ac |
| rs13121833 (4:1357358;C) | *FOXD1, FOXP1, FOXQ1* | — |
| rs2659510<br>(4:102113048;C) | *TFAP2A, TFAP2B, TFAB2C, MAFK, NRL* | — |
| rs26719 (5:92136381;T) | *LIN54* | — |
| rs36039219<br>(7:11704538;A) | *CREB1, PAX2, JUND, MEIS1, MEIS2, MEIS3, TBX2, TBX4,*<br>*TBX5* | — |
| rs10810957<br>(9:18332000;C) | *FOXD3, ARID5A, ZBTB1B* | — |
| rs11631291<br>(15:71889320;A) | *CREB1, KLF1, KLF4, KLF5, FOS:JUN* | H3K4me3 |
| rs12905641<br>(15:78964362;T) | *HAND1:TCF3, REL, RELA, STAT1, STAT3, STAT4, STAT5B,*<br>*STAT6, THAP1, NFATC1, NFATC3* | H3K4me1, H3K27ac |
| rs1885986 (17:2203175;G) | *ZNF354C, ZFX, ZBTB7C* | H3K36me3 |
| rs10423094<br>(19:49533380;T) | *USF1, USF2, TCF3, HEY2, SNAI2, FIGLA, ID4, TFEC*<br>*MLX1PL* | — |
| rs6082296<br>(20:21026509;C) | *Mafb, HNF1B, MEIS1, MEIS2, EVX1, EVX2, DUC, RHOX11* | H3K27me3, H3K4me1 |

#Active histone modification: H3K4me3(promoter-associated), H3K4me1(enhancer-associated), H3K9ac(promoter-associated), H3K27ac(enhancer/promoter-associated) H3K36me3(transcription-associated); Repressive histone modification: H3K27me3, H3K9me3

### Mapping of SNPs to genes

Mapping of the 15 SNPs to genes performed using FUMA identified a total of 122 genes in their vicinity (Table 4 and Supplementary Table 7). No protein coding genes were found around 20kb from three SNPs (rs1868172, rs26719, rs10810957). All the SNPs except for rs2659510 (4:102113048:A>C) were found to either interfere with expression or to interact with more than one gene through 3D chromatin interaction. Protein class analysis showed that most of the genes code for regulatory proteins such as those involved in chromatin modifier, transcription regulator and cellular signalling (Figure 2). Except for CGB1, all the others were found to have direct association with one or more phenotypes related to cognition, autism, schizophrenia and/or bipolar disorder (Supplementary Table 9). Based on the input criteria detailed in the methodology, VarElect showed highest score for *PAFAH1B1* followed by *FOXG1, FOS, FOXP2, TMEM106B, IDUA, CREB1, CHRNA3, CHRNA4PPP3CA* (Supplementary Table 10). Two most notable findings which emerged from this mapping effort and finctional annotation were i) Observing that of these 122 genes are either previously implicated candidate genes or are functionally relevant; and ii) 51 of these 122 genes and 79 TFs are druggable according to DGIB (https://www.dgidb.org/), a database curated from 31 sources such as DTC, DrugBank, TTD, and PharmaGKB. Many of these genes, such as, *CHRNA3, 4, HTR6* etc. are already being targeted for schizophrenia and psychosis by medications already in common use (clozapine, olanzapine, doxepin etc) or proposed for repurposing in these conditions (Supplementary Table 11).

**Table 4:**
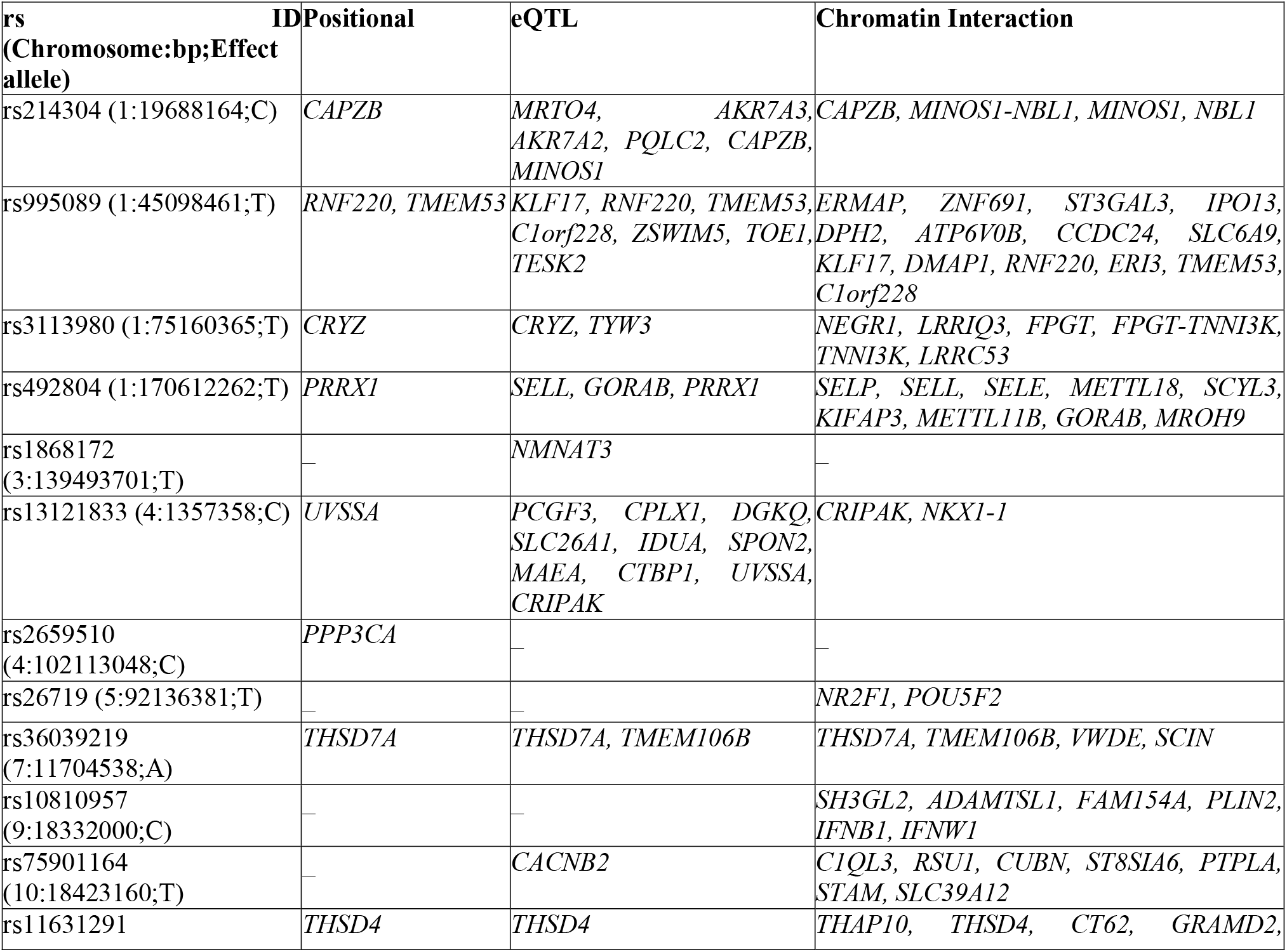
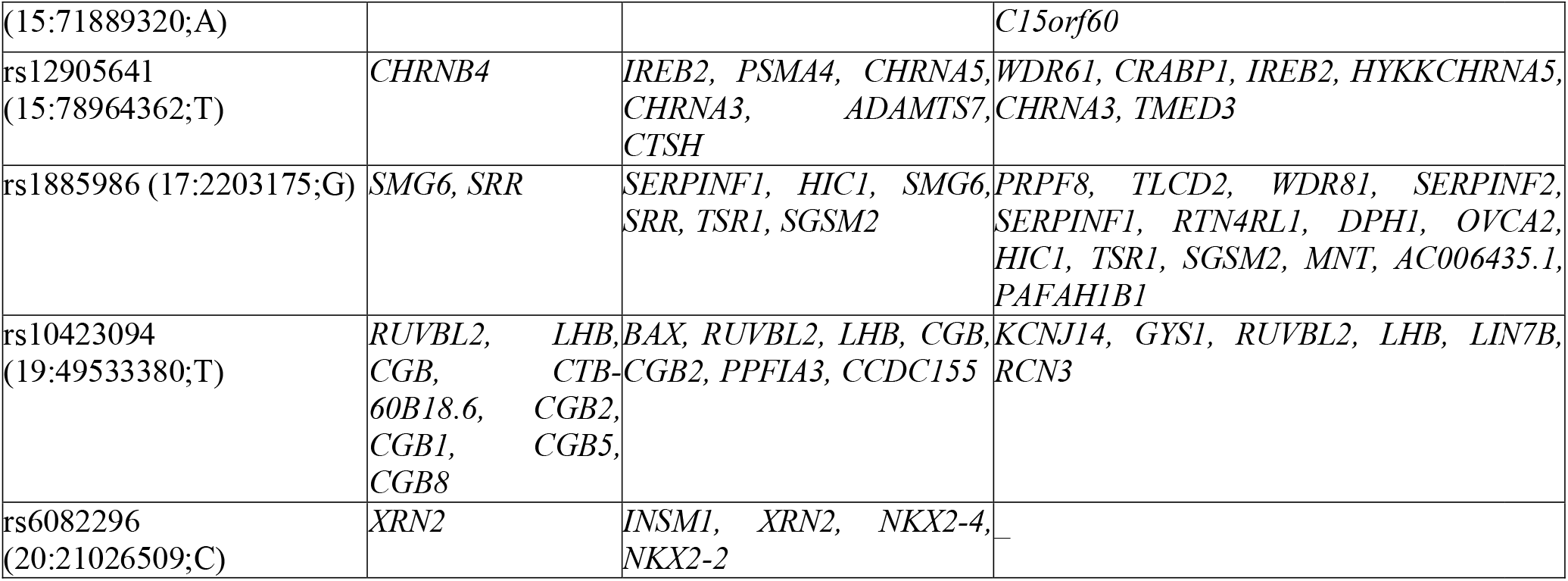
Mapping of associated SNPs to genes.

**Figure 2:**
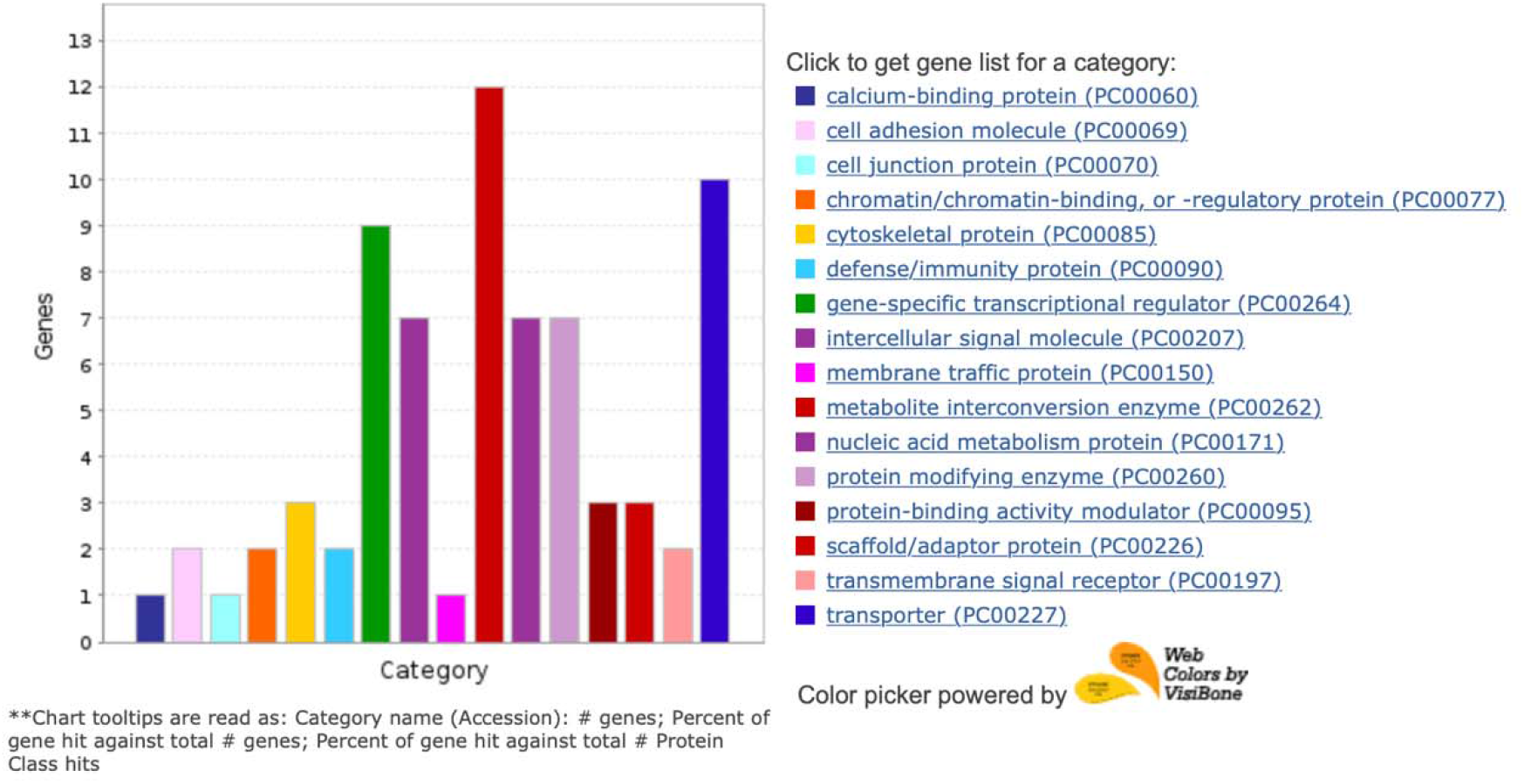
Protein class analysis of mapped genes.

### Pathway enrichment and functional annotation of the mapped genes and transcription factors

In the pathway enrichment analysis for 201 genes (122 mapped genes and 79 transcription factors whose binding sites were observed to be modulated by the associated SNPs; Table 3; Supplementary Table 8), PDGFR, EFG-EGFR, FGF and chemokine signaling pathways were found to be enriched. The top-most significantly enriched pathways relevant to cognition are shown in Table 5.

**Table 5:** Pathway enrichment of the mapped genes and transcription factors.

| Term | Adjusted P-value | Resource |
| --- | --- | --- |
| Th1 and Th2 cell differentiation | 7.82E-09 | KEGG_human_2019 |
| Hepatitis B | 7.82E-09 | KEGG_human_2019 |
| Th17 cell differentiation | 7.82E-09 | KEGG_human_2019 |
| Prolactin signalling pathway | 2.41E-05 | KEGG_human_2019 |
| Signalling pathways regulating pluripotency of stem cells | 3.77E-04 | KEGG_human_2019 |
| IL12-mediated signalling events | 1.92E-07 | NCI-Nature_2016 |
| IL2-mediated signalling events | 1.95E-04 | NCI-Nature_2016 |
| IL12 signalling mediated by STAT4 | 6.79E-06 | NCI-Nature_2016 |
| IL23-mediated signalling events | 6.49E-04 | NCI-Nature_2016 |
| Calcineurin-regulated Nuclear factor of activated T cells (NFAT)-dependent transcription in lymphocytes | 8.49E-05 | NCI-Nature_2016 |
| GMCSF-mediated signalling events Homo sapiens | 2.52E-04 | NCI-Nature_2016 |
| Glucocorticoid receptor regulatory network | 3.20E-08 | NCI-Nature_2016, Panther_2016 |
| Regulation of nuclear SMAD2/3 signalling Homo sapiens | 3.20E-08 | NCI-Nature_2016, Panther_2016 |
| IL27-mediated signaling events | 7.76E-04 | Panther2016 |
| PDGFR-beta pathway | 1.48E-06 | Wiki_pathway_human 2019 |
| VEGFA-VEGFR2 Signalling Pathway | 1.83E-05 | Wiki_pathway_human 2019 |
| IL-2 Signalling Pathway | 9.15E-06 | Wiki_pathway_human 2019 |
| IL-5 Signalling Pathway | 8.04E-06 | Wiki_pathway_human 2019 |
| IL-9 Signalling Pathway WP22 | 2.61E-04 | Wiki_pathway_human 2019 |
| IL-4 Signalling Pathway WP395 | 2.09E-05 | Wiki_pathway_human 2019 |
| EGF/EGFR Signalling Pathway | 1.83E-05 | Wiki_pathway_human 2019 |
| AGE/RAGE pathway | 1.29E-05 | Wiki_pathway_human 2019 |
| Brain-Derived Neurotrophic Factor (BDNF) signalling pathway | 2.13E-04 | Wiki_pathway_human 2019 |
| Interferon type I signalling pathways | 2.13E-04 | Wiki_pathway_human 2019 |
| Prolactin Signalling Pathway | 1.74E-04 | Wiki_pathway_human 2019 |

### Associated HPO phenotypes with the mapped genes and transcription factors

All the genes identified in this study were found to be associated with human phenotypes related to cognition such as abnormality of higher mental function, intellectual disability, abnormality of the nervous system, morphological abnormality of brain and nervous system (Table 6).

**Table 6:** HPO phenotypes associated with the mapped genes and transcription factors.

| HPO-Term | Overlapped genes/total genes<br>(P-value) |
| --- | --- |
| Abnormality of the nervous system (HP:0000707) | 54/3276 (6.237e-07) |
| Morphological central nervous system abnormality (HP:0002011) | 42/2307 (1.279e-06) |
| Abnormal nervous system morphology (HP:0012639) | 46/2504 (2.701e-07) |
| Abnormality of higher mental function (HP:0011446) | 39/2211 (7.009e-06) |
| Abnormality of brain morphology (HP:0012443) | 37/2106 (1.375e-05) |
| Intellectual disability (HP:0001249) | 30/1628 (4.230e-05) |
| Abnormal nervous system physiology (HP:0012638) | 48/3053 (1.155e-05) |

## Discussion

In comparison to other highly developed and intelligent great apes, such as the chimpanzees, the human brain is capable of supporting a wide range of complex cognitive abilities(Wei et al. 2019). This distinction is commonly believed to be associated with the evolution of the human brain(Maclean 2016; Sherwood, Subiaul, and Zawidzki 2008) which is driven by changes in gene regulation rather than divergence in protein-coding sequences(King and Wilson 1975). Thus, it is plausible to hypothesize that the development and evolution of the human brain could be an outcome of a discrete set of human-specific genetic alterations(Bae, Jayaraman, and Walsh 2015). Accordingly, HARs which evolved exclusively in humans and enriched in regulatory elements may drive evolution of human-specific traits via gene regulation(Won et al. 2019). Recent studies have identified HARs are associated with brain development (Bae, Jayaraman, and Walsh 2015), neuroanatomical or neurobehavioral phenotypes such as autism spectrum disorder(Doan et al. 2016), schizophrenia(Xu et al. 2015) and higher-order cognitive networks in the human brain(Wei et al. 2019). Thus, variants within HARs may alter the expression of genes under their regulation, which may result in a higher risk of cognitive impairment in schizophrenia patients.

Using linear regression analysis with disease status and HAR-variants as independent factors, principal components extracted from neurocognitive scores in eight different domains as dependent variables with age and gender as cofactors, we identified interaction affect six SNPs and disease status were significantly associated with the first two principal components (Table 2). These findings lend the first level of support to their likely contribution to the neurocognitive aspect of schizophrenia. Of note, four of these variants were common among the 13 variants that were observed to be associated with cognition in healthy control individuals (Table 2). The second notable support to the likely contribution of HAR variants to cognitive features/skills comes from the association of 12 of these SNPs in large GWASs of general cognitive function, educational attainment, neuroticism and/or intelligence (Supplementary Table 5). Though these were nominal associations, these GWAS cohorts could be considered as replication samples, and it is even more impressive given that these cohorts are mostly transethnic. Of note, rs1868172 showed suggestive genome wide significance (P-value 2.33E-05) with intelligence and was also found to be associated with other neurocognitive functions.

It may be further supportive to mention here that of the remaining 32 variants, 10 were nominally associated with one or more related phenotypes (Supplementary Table 12). Interestingly four HAR variants that showed association with schizophrenia in our previous study (Bhattacharyya et al. 2020) also showed significant association in several cognitive function GWAS; with rs3800926 strongly associated (P-value 6.1E-06) in a meta general cognition GWAS (Supplementary Table 12). This particular SNP showed nominal association across all GWASs of related traits, namely neurocortism, intelligence, and educational attainment (Supplementary Table 12). Neuroticism is characterized by chronic negative affect and susceptibility to stress, may be a risk factor for cognitive dysfunction(L. L. Boyle et al. 2010) while educational attainment and intelligence has been used as a proxy to cognitive function(Comes et al. 2019). These associations may not have been captured in either control or case-control cohort, due to limited sample size in our study. Nevertheless, these findings reiterate the contribution of HARs to higher order human cognitive abilities.

### Functional mapping

Functional mapping of these 15 associated variants revealed that these might affect binding of a total of 79 TF and/or overlap with active promoters, repressors or enhancers (Table 3, Supplementary Table 6). Through positional, eQTL and chromatin interaction mapping these SNPs were mapped to a total of 122 genes (Supplementary Table 7). Gene ontology analysis shows that most of the mapped genes are regulatory in nature, such as transcription regulator, chromatin modifier, protein modifier and binding regulator etc (Figure 2). Interestingly most of these mapped genes and TFs are implicated in neurocognitive phenotypes and diseases that have cognitive impairment as an endophenotype, such as, autism, schizophrenia and bipolar disorders (Supplementary Table 10). It is likely that genomic regions or genes near these 15 associated SNPs are also mostly regulatory in function and thus might have an important role in cognitive behaviour through their quantitative differences. Within the mapped genes and TFs whose binding sites are predicted altered by the HAR SNPs, the topmost genes according to Varelect scoring are *PAFAH1B1, TMEM106B, FOXP1, PPP3CA* etc (Supplementary Table 10), which have been previously implicated in general cognitive functions and diseases related to cognitive impairment.

Pathway enrichment analysis: The most significant pathways were chemokine mediated signalling pathways such as IL2, 4,5 9,12, 23, interferon type 1 and other immune response related pathways such as Th1/Th2/Th17 cell differentiation, NFAT dependent Ca 2+/calcineurin-regulated transcription in T cells and SMAD2/3 signalling; signalling pathways including BDNF, VEGFA-VEGFR2 etc which are implicated in brain development. New research suggests that stress-induced hippocampal neurogenesis alteration is accompanied by Th1/Th2 balance(Qi 2015). The central nervous system is not just regulated by a few specific cytokines, but rather by many cytokines, which are modulated by a systemic Th1/Th2 cytokine balance, which is associated with modulation of sophisticated brain functions(Qi 2015; Sredni-Kenigsbuch 2002). SMAD is generally known to elicit an inflammatory immune response when encountered with pathogens (Malhotra and Kang 2013). Recent research has demonstrated that SMAD2/3 can also maintain brain plasticity(Tecalco-Cruz et al. 2018). Several SMAD-binding proteins (e.g. Ski, SnoN) effectively influence neural development mainly through regulating SMAD activity(Tecalco-Cruz et al. 2018; Ueberham and Arendt 2013). Another immune response gene, Platelet-derived growth factor (PDGF) and its receptors (PDGFRs) are expressed in a number of cells, including neurons, astrocytes, and oligodendrocytes(Sil et al. 2018; Funa and Sasahara 2014). It is well-documented that PDGF-mediated signaling regulates a variety of central nervous system (CNS) functions including neurogenesis, cell survival, synaptogenesis and the differentiation of specific types of neurons(Sil et al. 2018; Funa and Sasahara 2014). Cytokines that include the interleukins (IL), interferons, tumor necrosis factors (TNF-α,β), and tumor growth factors, are released during neuronal activity and play a crucial role in regulating the strength of synaptic transmission (Bourgognon and Cavanagh 2020; Mousa and Bakhiet 2013; Prieto and Cotman 2017). Considering that cytokines are either proinflammatory (e.g., IL-6, IL-1β, TNF-α) or anti-inflammatory (e.g., IL-4, IL-10, IL-13) and are able to self-regulate immune response, ideally a balance of these two classes must exist to maintain optimal immune system function(Ray 2016; Cicchese et al. 2018). Immune response is one of the functions that have been more strongly targeted by natural selection during human evolution(Barreiro and Quintana-Murci 2010). The evolutionary genetic dissection of the immune system has greatly helped to distinguish genes and functions that are essential, redundant or advantageous for human survival. While there is a significant similarity in the coding sequences of cytokines and other immune response genes between humans and chimpanzees, the expression, signaling and immune response is quite different between these two species(Barreiro et al. 2010; Brinkworth and Babbitt 2019). It might be possible that HARs regulate the human specific gene expression and thus signaling of the nearby cytokine(s) or other immune response related genes.

At this point, in the context of schizophrenia or any other neuropsychiatric disorder, a co-ordination between immune response and nervous system needs some discussion. Nervous system and the immune system are two main regulators of homeostasis in the body that ensures normal organismal functioning and an effective communication between these is necessary. While CNS and microglia constantly monitor synapses and participate in their pruning during development and possibly also throughout life, immune cells and molecules sculpt the circuitry and modulate the activity of the nervous system. There is substantial evidence that innate immunity-related molecules, such as cytokines, toll-like receptors, the complement system, and acquired immunity-related molecules are expressed in the brain and play an important role in brain development(Marin and Kipnis 2013; Bilbo and Schwarz 2012; Morimoto and Nakajima 2019; Bennett and Molofsky 2019). Disruption of the immune system can manifest in cognitive declines and in neurodevelopmental abnormalities (Marin and Kipnis 2013; Bilbo and Schwarz 2012; Morimoto and Nakajima 2019; Bennett and Molofsky 2019). Inflammation within the brain is known to have far-reaching acute and long-term effects. In general, inflammation can produce a large number of proinflammatory factors (Russo et al., 2010) which can be harmful to brain cells and can have deleterious consequences for neuronal and cognitive function. A previous study showing that a significant number of HAR genes are targets of FDA-approved drugs, many of which are currently being used to treat neurological disorders underscore their importance in drug targeting and development of new therapeutics (Chu, Quan, and Zhang 2020). Although our sample size is too small to conclude causality, significant association of these associated SNPs in the large GWASs screened shows that these SNPs might have an impact neurocognitive function. These findings highlight the role of HAR-variants in cognition or related phenotypes and may lead to new or repurposed therapeutics that could help patients with impaired cognition and thus witness a more successful outcome.

In summary, our findings suggest that HAR variants are likely regulatory in nature and alter expression of genes in their vicinity through altered TF binding or chromatin structure. These consequent quantitative changes may confer higher risk of cognitive impairment in schizophrenia patients. Genes to which these associated SNPs mapped are known for their roles in biology of neurocognitive function as well as for their druggable nature as mentioned in the results section (Supplementary Table 11) lending support to the observed association and early inferences made in this study. Though very preliminary, these findings suggests that further analysis of HARs in general cognitive ability may enable i) better understanding of the role of HARs in human cognition; and ii) identify new drug targets and more targeted use of repurposed drugs to meet the continuing challenge of poor functional outcome in schizophrenia subjects due to diminished cognitive ability along with disease progression.

## Data Availability

Please see PENNCNB for cognitive scores of cases and controls

https://penncnp.med.upenn.edu

## Notes

### Competing Interest Statement

The authors have declared no competing interest.

### Funding Statement

ICMR
SERB
DST

### Author Declarations

Institutional Ethical committee of Ram Manohar Lohia Hospital

## References

1. Avila, Matthew, Gunvant Thaker, and Helene Adami. 2001. “Genetic Epidemiology and Schizophrenia: A Study of Reproductive Fitness.” Schizophrenia Research 47 (2–3): 233–41. https://doi.org/10.1016/S0920-9964(00)00062-1.

2. Bae, Byoung Il, Divya Jayaraman, and Christopher A. Walsh. 2015. “Genetic Changes Shaping the Human Brain.” Developmental Cell. Cell Press. https://doi.org/10.1016/j.devcel.2015.01.035.

3. Barkley, Russell A. 2001. “The Executive Functions and Self-Regulation: An Evolutionary Neuropsychological Perspective.” Neuropsychology Review. Springer. https://doi.org/10.1023/A:1009085417776.

4. Barreiro, Luis B., John C. Marioni, Ran Blekhman, Matthew Stephens, and Yoav Gilad. 2010. “Functional Comparison of Innate Immune Signaling Pathways in Primates.” Edited by Greg Gibson. PLoS Genetics 6 (12): e1001249. https://doi.org/10.1371/journal.pgen.1001249.

5. Barreiro, Luis B., and Lluís Quintana-Murci. 2010. “From Evolutionary Genetics to Human Immunology: How Selection Shapes Host Defence Genes.” Nature Reviews Genetics. Nat Rev Genet. https://doi.org/10.1038/nrg2698.

6. Bennett, F. C., and A. V. Molofsky. 2019. “The Immune System and Psychiatric Disease: A Basic Science Perspective.” Clinical & Experimental Immunology 197 (3): pcei.13334. https://doi.org/10.1111/cei.13334.

7. Bhatia, Triptish, Akhilesh Agarwal, Gyandeepak Shah, Joel Wood, Jan Richard, Raquel E. Gur, Ruben C. Gur, Vishwajit L. Nimgaonkar, Sati Mazumdar, and Smita N. Deshpande. 2012. “Adjunctive Cognitive Remediation for Schizophrenia Using Yoga: An Open, Non-Randomised Trial.” Acta Neuropsychiatrica 24 (02): 91–100. https://doi.org/10.1111/j.1601-5215.2011.00587.x.

8. Bhattacharyya, Upasana, Smita N Deshpande, Triptish Bhatia, and B K Thelma. 2020. “Revisiting Schizophrenia from an Evolutionary Perspective: An Association Study of Recent Evolutionary Markers and Schizophrenia.” Schizophrenia Bulletin, December. https://doi.org/10.1093/schbul/sbaa179.

9. Bilbo, Staci D., and Jaclyn M. Schwarz. 2012. “The Immune System and Developmental Programming of Brain and Behavior.” Frontiers in Neuroendocrinology. NIH Public Access. https://doi.org/10.1016/j.yfrne.2012.08.006.

10. Bourgognon, Julie-Myrtille, and Jonathan Cavanagh. 2020. “The Role of Cytokines in Modulating Learning and Memory and Brain Plasticity.” Brain and Neuroscience Advances 4 (January): 239821282097980. https://doi.org/10.1177/2398212820979802.

11. Bowie, Christopher R., and Philip D. Harvey. 2006. “Cognitive Deficits and Functional Outcome in Schizophrenia.” Neuropsychiatric Disease and Treatment. Dove Press. https://doi.org/10.2147/nedt.2006.2.4.531.

12. Boyle, Alan P., Eurie L. Hong, Manoj Hariharan, Yong Cheng, Marc A. Schaub, Maya Kasowski, Konrad J. Karczewski, et al. 2012. “Annotation of Functional Variation in Personal Genomes Using RegulomeDB.” Genome Research 22 (9): 1790–97. https://doi.org/10.1101/gr.137323.112.

13. Boyle, Lisa L., Jeffrey M. Lyness, Paul R. Duberstein, Jurgis Karuza, Deborah A. King, Susan Messing, and Xin Tu. 2010. “Trait Neuroticism, Depression, and Cognitive Function in Older Primary Care Patients.” American Journal of Geriatric Psychiatry 18 (4): 305–12. https://doi.org/10.1097/JGP.0b013e3181c2941b.

14. Brinkworth, Jessica F, and Courtney C Babbitt. 2019. “Immune System Promiscuity in Human and Non-Human Primate Evolution.” https://digitalcommons.wayne.edu/humbiol_preprints/143.

15. Calafato, Maria Stella, and Elvira Bramon. 2019. “Interplay between Genetics, Cognition and Schizophrenia | Brain | Oxford Academic.” Brain 142 (2): 236–38. https://academic.oup.com/brain/article/142/2/236/5304056.

16. Capra, John A., Genevieve D. Erwin, Gabriel McKinsey, John L. R. Rubenstein, and Katherine S. Pollard. 2013a. “Many Human Accelerated Regions Are Developmental Enhancers.” Philosophical Transactions of the Royal Society B: Biological Sciences 368 (1632): 20130025. https://doi.org/10.1098/rstb.2013.0025.

17. ———. 2013b. “Many Human Accelerated Regions Are Developmental Enhancers.” Philosophical Transactions of the Royal Society B: Biological Sciences 368 (1632): 20130025. https://doi.org/10.1098/rstb.2013.0025.

18. Capra, John A., Genevieve D. Erwin, Gabriel Mckinsey, John L.R. Rubenstein, and Katherine S. Pollard. 2013. “Many Human Accelerated Regions Are Developmental Enhancers.” Philosophical Transactions of the Royal Society B: Biological Sciences 368 (1632): 20130025. https://doi.org/10.1098/rstb.2013.0025.

19. Carroll, Sean B. 2005. “Evolution at Two Levels: On Genes and Form.” PLoS Biology 3 (7): e245. https://doi.org/10.1371/journal.pbio.0030245.

20. Chen, Edward Y., Christopher M. Tan, Yan Kou, Qiaonan Duan, Zichen Wang, Gabriela V. Meirelles, Neil R. Clark, and Avi Ma’ayan. 2013. “Enrichr: Interactive and Collaborative HTML5 Gene List Enrichment Analysis Tool.” BMC Bioinformatics 14 (nApril). https://doi.org/10.1186/1471-2105-14-128.

21. Chu, Xin Yi, Yuan Quan, and Hong Yu Zhang. 2020. “Human Accelerated Genome Regions with Value in Medical Genetics and Drug Discovery.” Drug Discovery Today. Elsevier Ltd. https://doi.org/10.1016/j.drudis.2020.03.001.

22. Cicchese, Joseph M., Stephanie Evans, Caitlin Hult, Louis R. Joslyn, Timothy Wessler, Jess A. Millar, Simeone Marino, et al. 2018. “Dynamic Balance of Pro- and Anti-Inflammatory Signals Controls Disease and Limits Pathology.” Immunological Reviews 285 (1): 147–67. https://doi.org/10.1111/imr.12671.

23. Comes, Ashley L., Fanny Senner, Monika Budde, Kristina Adorjan, Heike Anderson-Schmidt, Till F.M. Andlauer, Katrin Gade, et al. 2019. “The Genetic Relationship between Educational Attainment and Cognitive Performance in Major Psychiatric Disorders.” Translational Psychiatry 9 (1): 1–11. https://doi.org/10.1038/s41398-019-0547-x.

24. Crow, T. J. 2000. “Schizophrenia as the Price That Homo Sapiens Pays for Language: A Resolution of the Central Paradox in the Origin of the Species.” In Brain Research Reviews, 31:118–29. https://doi.org/10.1016/S0165-0173(99)00029-6.

25. Crow, Timothy J. 1997. “Is Schizophrenia the Price That Homo Sapiens Pays for Language?” In Schizophrenia Research, 28:127–41. https://doi.org/10.1016/S0920-9964(97)00110-2.

26. d’Errico, Francesco, Christopher Henshilwood, Graeme Lawson, Marian Vanhaeren, Anne Marie Tillier, Marie Soressi, Frédérique Bresson, et al. 2003. “Archaeological Evidence for the Emergence of Language, Symbolism, and Music - An Alternative Multidisciplinary Perspective.” Journal of World Prehistory 17 (1): 1–70. https://doi.org/10.1023/A:1023980201043.

27. Davies, G., R. E. Marioni, D. C. Liewald, W. D. Hill, S. P. Hagenaars, S. E. Harris, S. J. Ritchie, et al. 2016. “Genome-Wide Association Study of Cognitive Functions and Educational Attainment in UK Biobank (N=112 151).” Molecular Psychiatry 21 (6): 758–67. https://doi.org/10.1038/mp.2016.45.

28. Davies, Gail, Max Lam, Sarah E. Harris, Joey W. Trampush, Michelle Luciano, W. David Hill, Saskia P. Hagenaars, et al. 2018. “Study of 300,486 Individuals Identifies 148 Independent Genetic Loci Influencing General Cognitive Function.” Nature Communications 9 (1): 1–16. https://doi.org/10.1038/s41467-018-04362-x.

29. Deshpande, Smita N., Mohit N.L. Mathur, Shri K. Das, Triptish Bhatia, Shridhar Sharma, and Vishwajit L. Nimgaonkar. 1998. “A Hindi Version of the Diagnostic Interview for Genetic Studies.” Schizophrenia Bulletin 24 (3): 489–93. https://doi.org/10.1093/oxfordjournals.schbul.a033343.

30. Doan, Ryan N., Byoung Il Bae, Beatriz Cubelos, Cindy Chang, Amer A. Hossain, Samira Al-Saad, Nahit M. Mukaddes, et al. 2016. “Mutations in Human Accelerated Regions Disrupt Cognition and Social Behavior.” Cell 167 (2): 341-354.e12. https://doi.org/10.1016/j.cell.2016.08.071.

31. Dongen, Jenny Van, and Dorret I Boomsma. n.d. “The Evolutionary Paradox and the Missing Heritability of Schizophrenia.” https://doi.org/10.1002/ajmg.b.32135.

32. Emre, Bora, Yücel Murat, and Pantelis Christos. 2010. “Cognitive Impairment in Schizophrenia and Affective Psychoses: Implications for DSM-V Criteria and Beyond | Schizophrenia Bulletin | Oxford Academic.” Schizophrenia Bulletin 36 (1): 36–42. https://academic.oup.com/schizophreniabulletin/article/36/1/36/1868817.

33. Fox, John, and Sanford Weisberg. n.d. “An R Companion to Applied Regression | SAGE Publications Inc.” Accessed January 9, 2021. <https://us.sagepub.com/en-us/nam/an-r-companion-to-applied-regression/book246125>.

34. Funa, Keiko, and Masakiyo Sasahara. 2014. “The Roles of PDGF in Development and during Neurogenesis in the Normal and Diseased Nervous System.” Journal of Neuroimmune Pharmacology. Springer New York LLC. https://doi.org/10.1007/s11481-013-9479-z.

35. Green, M. F., R. S. Kern, D. L. Braff, and J. Mintz. 2000. “Neurocognitive Deficits and Functional Outcome in Schizophrenia: Are We Measuring the ‘Right Stuff’?” Schizophrenia Bulletin 26 (1): 119–36. https://doi.org/10.1093/oxfordjournals.schbul.a033430.

36. Harvey, Philip D. n.d. “The Genetics of Cognitive Impairment in Schizophrenia.” Psychiatry (Edgmont) 5 (6): 65.

37. Hawkins, D. M., and Sanford Weisberg. n.d. “Sabinet | Combining the Box-Cox Power and Generalised Log Transformations to Accommodate Nonpositive Responses in Linear and Mixed-Effects Linear Models.” Accessed January 9, 2021. https://journals.co.za/content/journal/10520/EJC-bd05f9440.

38. Hellard Stéphanie Le, Yunpeng Wang, Aree Witoelar, Verena Zuber, Francesco Bettella, Kenneth Hugdahl, Thomas Espeseth, et al. 2016. “Identification of Gene Loci That Overlap Between Schizophrenia and Educational Attainment.” Schizophrenia Bulletin 43 (3): sbw085. https://doi.org/10.1093/schbul/sbw085.

39. Heuvel, Martijn P. Van Den, Lianne H. Scholtens, Siemon C. De Lange, Rory Pijnenburg, Wiepke Cahn, Neeltje E.M. Van Haren, Iris E. Sommer, et al. 2019. “Evolutionary Modifications in Human Brain Connectivity Associated with Schizophrenia.” Brain 142 (12): 3991–4002. https://doi.org/10.1093/brain/awz330.

40. Heyes, Cecilia. 2012. “New Thinking: The Evolution of Human Cognition.” Philosophical Transactions of the Royal Society B: Biological Sciences 367 (1599): 2091–96. https://doi.org/10.1098/rstb.2012.0111.

41. Hori, Hiroaki, Noriko Yamamoto, Takashi Fujii, Toshiya Teraishi, Daimei Sasayama, Junko Matsuo, Yumiko Kawamoto, et al. 2012. “Effects of the CACNA1C Risk Allele on Neurocognition in Patients with Schizophrenia and Healthy Individuals.” Scientific Reports 2. https://doi.org/10.1038/srep00634.

42. Hubbard, Leon, Katherine E Tansey, Dheeraj Rai, Peter Jones, Stephan Ripke, Kimberly D Chambert, Jennifer L Moran, et al. 2016. “Evidence of Common Genetic Overlap Between Schizophrenia and Cognition.” Schizophrenia Bulletin 42 (3): 832–42. https://doi.org/10.1093/schbul/sbv168.

43. Hubisz, Melissa J., and Katherine S. Pollard. 2014. “Exploring the Genesis and Functions of Human Accelerated Regions Sheds Light on Their Role in Human Evolution.” Current Opinion in Genetics and Development. Elsevier Ltd. https://doi.org/10.1016/j.gde.2014.07.005.

44. Kamm, Gretel B., Francisco Pisciottano, Rafi Kliger, and Lucía F. Franchini. 2013. “The Developmental Brain Gene NPAS3 Contains the Largest Number of Accelerated Regulatory Sequences in the Human Genome.” Molecular Biology and Evolution 30 (5): 1088–1102. https://doi.org/10.1093/molbev/mst023.

45. King, MC, and AC Wilson. 1975. “Evolution at Two Levels in Humans and Chimpanzees.” Science 188.

46. Kukshal, Prachi, Triptish Bhatia, A. M. Bhagwat, Raquel E. Gur, Ruben C. Gur, Smita N. Deshpande, Vishwajit L. Nimgaonkar, and B. K. Thelma. 2013a. “Association Study of Neuregulin-1 Gene Polymorphisms in a North Indian Schizophrenia Sample.” Schizophrenia Research 144 (1–3): 24–30. https://doi.org/10.1016/j.schres.2012.12.017.

47. Kukshal, Prachi, Triptish Bhatia, A.M. Bhagwat, Raquel E. Gur, Ruben C. Gur, Smita N. Deshpande, Vishwajit L. Nimgaonkar, and B.K. Thelma. 2013b. “Association Study of Neuregulin-1 Gene Polymorphisms in a North Indian Schizophrenia Sample.” Schizophrenia Research 144 (1–3): 24–30. https://doi.org/10.1016/j.schres.2012.12.017.

48. Kuleshov MV, Jones MR, Rouillard AD, Fernandez NF, Duan Q, Wang Z, Koplev S, Jenkins SL, Jagodnik KM, Lachmann A, McDermott MG, Monteiro CD, Gundersen GW, Ma’ayan A. 2016. “Enrichr: A Comprehensive Gene Set Enrichment Analysis Web Server 2016 Update - PubMed.” Nucleic Acids Research, 44(W1)-W90–7. https://pubmed.ncbi.nlm.nih.gov/27141961/?from_single_result=Enrichr%3A+A+Comprehensive+Gene+Set+Enrichment+Analysis+Web+Server+2016+Update+expanded_search_query=Enrichr%3A+A+Comprehensive+Gene+Set+Enrichment+Analysis+Web+Server+2016+Update.

49. Lee, James J., Robbee Wedow, Aysu Okbay, Edward Kong, Omeed Maghzian, Meghan Zacher, Tuan Anh Nguyen-Viet, et al. 2018. “Gene Discovery and Polygenic Prediction from a Genome-Wide Association Study of Educational Attainment in 1.1 Million Individuals.” Nature Genetics 50 (8): 1112–21. https://doi.org/10.1038/s41588-018-0147-3.

50. Liu, Chenxing, Ian Everall, Christos Pantelis, and Chad Bousman. 2019. “Interrogating the Evolutionary Paradox of Schizophrenia: A Novel Framework and Evidence Supporting Recent Negative Selection of Schizophrenia Risk Alleles.” Frontiers in Genetics 10 (APR): 389. https://doi.org/10.3389/fgene.2019.00389.

51. Luciano, Michelle, Saskia P. Hagenaars, Gail Davies, W. David Hill, Toni Kim Clarke, Masoud Shirali, Sarah E. Harris, et al. 2018. “Association Analysis in over 329,000 Individuals Identifies 116 Independent Variants Influencing Neuroticism.” Nature Genetics 50 (1): 6–11. https://doi.org/10.1038/s41588-017-0013-8.

52. Ma, Chun-yu, Pamela Madden, Paul Gontarz, Ting Wang, and Bo Zhang. 2019. “FeatSNP: An Interactive Database for Brain-Specific Epigenetic Annotation of Human SNPs.” Frontiers in Genetics 10 (April): 262. https://doi.org/10.3389/fgene.2019.00262.

53. Maclean, Evan L. 2016. “Unraveling the Evolution of Uniquely Human Cognition.” Proceedings of the National Academy of Sciences of the United States of America 113 (23): 6348–54. https://doi.org/10.1073/pnas.1521270113.

54. Malhotra, Nidhi, and Joonsoo Kang. 2013. “SMAD Regulatory Networks Construct a Balanced Immune System.” Immunology 139 (1): 1–10. https://doi.org/10.1111/imm.12076.

55. Marin, Ioana, and Jonathan Kipnis. 2013. “Learning and Memory… and the Immune System.” Learning and Memory. Cold Spring Harbor Laboratory Press. https://doi.org/10.1101/lm.028357.112.

56. Morimoto, Keiko, and Kazunori Nakajima. 2019. “Role of the Immune System in the Development of the Central Nervous System.” Frontiers in Neuroscience. Frontiers Media S.A. https://doi.org/10.3389/fnins.2019.00916.

57. Mousa, Alyaa, and Moiz Bakhiet. 2013. “Role of Cytokine Signaling during Nervous System Development.” International Journal of Molecular Sciences. Multidisciplinary Digital Publishing Institute (MDPI). https://doi.org/10.3390/ijms140713931.

58. Nagel, M., D. Speed, S. van der Sluis, and S.D. Østergaard. 2020. “Genome-Wide Association Study of the Sensitivity to Environmental Stress and Adversity Neuroticism Cluster.” Acta Psychiatrica Scandinavica. Blackwell Publishing Ltd. https://doi.org/10.1111/acps.13155.

59. Nagel, Mats, Philip R. Jansen, Sven Stringer, Kyoko Watanabe, Christiaan A. De Leeuw, Julien Bryois, Jeanne E. Savage, et al. 2018. “Meta-Analysis of Genome-Wide Association Studies for Neuroticism in 449,484 Individuals Identifies Novel Genetic Loci and Pathways.” Nature Genetics 50 (7): 920–27. https://doi.org/10.1038/s41588-018-0151-7.

60. Nagel, Mats, Kyoko Watanabe, Sven Stringer, Danielle Posthuma, and Sophie Van Der Sluis. 2018. “Item-Level Analyses Reveal Genetic Heterogeneity in Neuroticism.” Nature Communications 9 (1): 1–10. https://doi.org/10.1038/s41467-018-03242-8.

61. Nurnberger, J. I., M. C. Blehar, C. A. Kaufmann, C. York-Cooler, S. G. Simpson, J. Harkavy-Friedman, J. B. Severe, et al. 1994. “Diagnostic Interview for Genetic Studies: Rationale, Unique Features, and Training.” Archives of General Psychiatry. American Medical Association. https://doi.org/10.1001/archpsyc.1994.03950110009002.

62. Ohi, Kazutaka, Chika Sumiyoshi, Haruo Fujino, Yuka Yasuda, Hidenaga Yamamori, Michiko Fujimoto, Tomoko Shiino, Tomiki Sumiyoshi, and Ryota Hashimoto. 2018. “Molecular Sciences Genetic Overlap between General Cognitive Function and Schizophrenia: A Review of Cognitive GWASs.” https://doi.org/10.3390/ijms19123822.

63. Okbay, Aysu, Bart M.L. Baselmans, Jan Emmanuel De Neve, Patrick Turley, Michel G. Nivard, Mark Alan Fontana, S. Fleur W. Meddens, et al. 2016. “Genetic Variants Associated with Subjective Well-Being, Depressive Symptoms, and Neuroticism Identified through Genome-Wide Analyses.” Nature Genetics 48 (6): 624–33. https://doi.org/10.1038/ng.3552.

64. Okbay, Aysu, Jonathan P. Beauchamp, Mark Alan Fontana, James J. Lee, Tune H. Pers, Cornelius A. Rietveld, Patrick Turley, et al. 2016. “Genome-Wide Association Study Identifies 74 Loci Associated with Educational Attainment.” Nature 533 (7604): 539–42. https://doi.org/10.1038/nature17671.

65. Pollard, Katherine S., Sofie R. Salama, Bryan King, Andrew D. Kern, Tim Dreszer, Sol Katzman, Adam Siepel, et al. 2006. “Forces Shaping the Fastest Evolving Regions in the Human Genome.” PLoS Genetics 2 (10): e168. https://doi.org/10.1371/journal.pgen.0020168.

66. Prabhakar, Shyam, Axel Visel, Jennifer A. Akiyama, Malak Shoukry, Keith D. Lewis, Amy Holt, Ingrid Plajzer-Frick, et al. 2008. “Human-Specific Gain of Function in a Developmental Enhancer.” Science 321 (5894): 1346–50. https://doi.org/10.1126/science.1159974.

67. Prieto, G. Aleph, and Carl W. Cotman. 2017. “Cytokines and Cytokine Networks Target Neurons to Modulate Long-Term Potentiation.” Cytokine and Growth Factor Reviews. Elsevier Ltd. https://doi.org/10.1016/j.cytogfr.2017.03.005.

68. Punchaichira, Toyanji Joseph, Prachi Kukshal, Triptish Bhatia, Smita Neelkanth Deshpande, and B. K. Thelma. 2020. “The Effect of Rs1076560 (DRD2) and Rs4680 (COMT) on Tardive Dyskinesia and Cognition in Schizophrenia Subjects.” Psychiatric Genetics, 125–35. https://doi.org/10.1097/YPG.0000000000000258.

69. Qi, Fangfang. 2015. “Immune-Based Modulation of Adult Hippocampal Neurogenesis, Link to Systemic Th1/Th2 Balance.” Journal of Vaccines & Vaccination 06 (02). https://doi.org/10.4172/2157-7560.1000274.

70. Ray, Arunabha. 2016. “Cytokines and Their Role in Health and Disease: A Brief Overview.” MOJ Immunology 4 (2). https://doi.org/10.15406/moji.2016.04.00121.

71. Rund, B. R. 1998. “A Review of Longitudinal Studies of Cognitive Functions in Schizophrenia Patients.” Schizophrenia Bulletin 24 (3): 425–35. https://doi.org/10.1093/oxfordjournals.schbul.a033337.

72. Ryu, Hane, Fumitaka Inoue, Sean Whalen, Alex Williams, Martin Kircher, Beth Martin, Beatriz Alvarado, et al. n.d. “Massively Parallel Dissection of Human Accelerated Regions in Human and Chimpanzee Neural Progenitors.” Accessed March 9, 2020. https://doi.org/10.1101/256313.

73. Savage, Jeanne E., Philip R. Jansen, Sven Stringer, Kyoko Watanabe, Julien Bryois, Christiaan A. De Leeuw, Mats Nagel, et al. 2018. “Genome-Wide Association Meta-Analysis in 269,867 Individuals Identifies New Genetic and Functional Links to Intelligence.” Nature Genetics 50 (7): 912–19. https://doi.org/10.1038/s41588-018-0152-6.

74. Sherwood, Chet C., Francys Subiaul, and Tadeusz W. Zawidzki. 2008. “A Natural History of the Human Mind: Tracing Evolutionary Changes in Brain and Cognition.” In Journal of Anatomy, 212:426–54. Wiley-Blackwell. https://doi.org/10.1111/j.1469-7580.2008.00868.x.

75. Sil, Susmita, Palsamy Periyasamy, Annadurai Thangaraj, Ernest T. Chivero, and Shilpa Buch. 2018. “PDGF/PDGFR Axis in the Neural Systems.” Molecular Aspects of Medicine. Elsevier Ltd. https://doi.org/10.1016/j.mam.2018.01.006.

76. Smeland, Olav B., and Ole A. Andreassen. 2018. “How Can Genetics Help Understand the Relationship between Cognitive Dysfunction and Schizophrenia?” Scandinavian Journal of Psychology 59 (1): 26–31. https://doi.org/10.1111/sjop.12407.

77. Sniekers, Suzanne, Sven Stringer, Kyoko Watanabe, Philip R. Jansen, Jonathan R.I. Coleman, Eva Krapohl, Erdogan Taskesen, et al. 2017. “Genome-Wide Association Meta-A Nalysis of 78,308 Individuals Identifies New Loci and Genes Influencing Human Intelligence.” Nature Genetics 49 (7): 1107–12. https://doi.org/10.1038/ng.3869.

78. Sredni-Kenigsbuch, Dvora. 2002. “Th1/Th2 Cytokines in the Central Nervous System.” International Journal of Neuroscience 112 (6): 665–703. https://doi.org/10.1080/00207450290025725.

79. Srinivasan, Saurabh, Francesco Bettella, Oleksandr Frei, W. David Hill, Yunpeng Wang, Aree Witoelar, Andrew J. Schork, et al. 2018. “Enrichment of Genetic Markers of Recent Human Evolution in Educational and Cognitive Traits.” Scientific Reports 8 (1). https://doi.org/10.1038/s41598-018-30387-9.

80. Srinivasan, Saurabh, Francesco Bettella, Morten Mattingsdal, Yunpeng Wang, Aree Witoelar, Andrew J. Schork, Wesley K. Thompson, et al. 2016. “Genetic Markers of Human Evolution Are Enriched in Schizophrenia.” Biological Psychiatry 80 (4): 284–92. https://doi.org/10.1016/j.biopsych.2015.10.009.

81. Tecalco-Cruz, Angeles C., Diana G. Ríos-López, Genaro Vázquez-Victorio, Reyna E. Rosales-Alvarez, and Marina Macías-Silva. 2018. “Transcriptional Cofactors Ski and SnoN Are Major Regulators of the TGF-β/Smad Signaling Pathway in Health and Disease.” Signal Transduction and Targeted Therapy. Springer Nature. https://doi.org/10.1038/s41392-018-0015-8.

82. Tomasello, Michael, and Esther Herrmann. 2010. “Ape and Human Cognition.” Current Directions in Psychological Science 19 (1): 3–8. https://doi.org/10.1177/0963721409359300.

83. Turley, Patrick, Raymond K. Walters, Omeed Maghzian, Aysu Okbay, James J. Lee, Mark Alan Fontana, Tuan Anh Nguyen-Viet, et al. 2018. “Multi-Trait Analysis of Genome-Wide Association Summary Statistics Using MTAG.” Nature Genetics 50 (2): 229–37. https://doi.org/10.1038/s41588-017-0009-4.

84. Ueberham, Uwe, and Thomas Arendt. 2013. “The Role of Smad Proteins for Development, Differentiation and Dedifferentiation of Neurons.” In Trends in Cell Signaling Pathways in Neuronal Fate Decision. InTech. https://doi.org/10.5772/54532.

85. Wei, Yongbin, Siemon C. de Lange, Lianne H. Scholtens, Kyoko Watanabe, Dirk Jan Ardesch, Philip R. Jansen, Jeanne E. Savage, et al. 2019. “Genetic Mapping and Evolutionary Analysis of Human-Expanded Cognitive Networks.” Nature Communications 10 (1): 1–11. https://doi.org/10.1038/s41467-019-12764-8.

86. Won, Hyejung, Jerry Huang, Carli K. Opland, Chris L. Hartl, and Daniel H. Geschwind. 2019. “Human Evolved Regulatory Elements Modulate Genes Involved in Cortical Expansion and Neurodevelopmental Disease Susceptibility.” Nature Communications 10 (1): 1–11. https://doi.org/10.1038/s41467-019-10248-3.

87. Xu, Ke, Eric E Schadt, Katherine S Pollard, Panos Roussos, and Joel T Dudley. 2015. “Genomic and Network Patterns of Schizophrenia Genetic Variation in Human Evolutionary Accelerated Regions.” Molecular Biology and Evolution 32 (5): 1148–60. https://doi.org/10.1093/molbev/msv031.

